# Exposition to airborne SARS-CoV-2 in household and hospitals settings as studied by a vacuum cleaner as a high-powered air sampler

**DOI:** 10.1101/2021.07.16.21260337

**Authors:** Peter de Man, Marco Ortiz, Philomena M. Bluyssen, Stijn J. de Man, Marie-Jozé Rentmeester, Marijke van der Vliet, Evert-Jan Wils, David S.Y. Ong

## Abstract

**Background:** We aimed to study the presence of SARS-CoV-2 in air surrounding infected healthcare workers (HCW) in their homes versus infected patients who were undergoing potential aerosol-generating medical procedures (AGMP). We also studied the effect of different face masks worn bij infected persons on spread of SARS-CoV-2 into the air.

**Methods:** We developed a high-volume air sampler method that uses a household vacuum cleaner with a surgical mask serving as a sample filter. SARS-CoV-2 RNA was harvested from these sample filters and analyzed on the presence of RNA by polymerase chain reaction. We acquired air samples in close aproximity of HCWs wearing different facemasks. Also, we obtained free air samples away from the infected HCWs and samples near intensive care unit (ICU) patients undergoing AGMP. Fog experiments were performed to visualize the airflow around our air sampler.

**Results:** Aerosols were visibly suctioned into the vacuum cleaner when there was no face mask, whereas wearing a face mask resulted in a delayed and reduced flow of aerosols into the vacuum cleaner. The face masks that were worn by the HCWs were positive in 54-83% of cases. The proportion of positive air samples was higher in household settings of recently infected HCWs (29/41; 70.7%) compared to ICU settings (4/17; 23.5%) (p<0.01).

**Conclusion:** This high-volume air sampler method was able to detect SARS-CoV-2 RNA in air samples. Air samples in the household environment of recently infected HCWs more frequently contained SARS-CoV-2 in comparison to those obtained in patient rooms during potential AGMP.

## Introduction

Severe acute respiratory syndrome coronavirus 2 (SARS-CoV-2) has three possible transmission routes: indirect contact transmission via deposited or transmitted infectious droplets via surfaces, direct transmission of virus-carrying droplets when in close vicinity, and airborne transmission through aerosols emitted by infected individuals.^1^ Since the start of the coronavirus disease 2019 (COVID-19) pandemic, both national and international healthcare authorities have heterogeneously valued the relative contributions and importance of each of these various transmission routes to the cumulative spread of COVID-19.^2–5^ Subsequently, determining the most important containment measure remains debatable as each measure is usually related to one specific transmission route more than to the other. Cleaning surfaces, washing hands, and sneezing/coughing in the elbow have been adopted to reduce both indirect and direct transmission. Physical distancing of individuals and wearing face masks are primarily aimed at preventing direct transmission from large infectious droplets, whereas adequate air ventilation would be relevant in the context of preventing airborne transmission.

Usually, mathematical models and observations from experimental or epidemiological investigations are used to assess the importance of the different transmission routes. However, real-life measurements on the presence of SARS-CoV-2 RNA in the air surrounding infected individuals would likely provide more accurate information. Previous studies used several types of air samplers based on different techniques, but show heterogeneous results with overall low yields, and underline the necessity for sampling larger air volumes because viruses are only present at very low concentrations in the air.^6^

We developed a high-volume air sampler method that uses a household vacuum cleaner with a surgical mask serving as a sample filter. This filter was tested on the presence of SARS-CoV-2 RNA by polymerase chain reaction (PCR). In this study, we aimed to estimate the efficacy of different face masks worn by infected persons on the spread of SARS-CoV-2 into the surrounding air in household settings and to assess the presence of SARS-CoV-2 in the air of private homes of infected individuals and in intensive care unit (ICU) rooms of patients who were undergoing different forms of potential aerosol-generating medical procedures (AGMP).

## Methods

### Study design

The study was performed at two sites between October 1, 2020, and January 22, 2021. First, air sampling experiments were performed in the private homes of SARS-CoV-2 positive healthcare workers (HCWs) while they wore different types of face masks. Second, air sampling was performed in ICU rooms of COVID-19 patients.

As part of hospital policy, HCWs were tested with combined throat nasopharyngeal swabs in case of symptoms suggestive of SARS-CoV-2 infection. We selected HCWs with positive SARS-CoV-2 results who had high virus loads (i.e., reverse transcriptase quantitative polymerase chain reaction (RT-qPCR) cycle threshold (Ct) values lower than 21), and performed the experiments in their homes within 24 hours after the positive test results.

Air samples in the ICU setting were collected in proximity (i.e., about 50 cm distance) of COVID-19 patients undergoing invasive mechanical ventilation (iMV), and AGMP such as high-flow nasal canulae (HFNC) therapy and intubation. Patients were selected irrespective of the virus load measured in the nasopharyngeal swabs. The ICU rooms had mechanical room ventilation with an air exchange rate of six times per hour.

The Institutional Review Board of the Franciscus Gasthuis and Vlietland Hospital in Rotterdam approved the study protocol and ethical approval was obtained (IRB protocol number 2020-092). Written informed consent was obtained from the HCWs during the household visits. As no specific instructions for mask-wearing or other behavior requirements were given to ICU patients in the second part of the study and only air samples were collected, the need for written informed consent was waived for this particular part of the study. The study was performed in accordance with the Helsinki Declaration as revised in 2013.

### Performance vacuum cleaner

Air sampling was performed using a Nilfisk household vacuum cleaner (model Elite performance comfort, 2000 watt), which has a HEPA filter on the air outlet. To assess the performance of our method, the volumetric airflow of the vacuum cleaner was measured with the Acin FlowFinder mk-2 in the SenseLab.^7^ The air velocity of the suction was measured with the DANTEC Dynamics ComfortSense air velocity meter. For the visualization experiment aerosols, with diameters ranging from 10 to 50 μm, were produced with polypropylene glycol with the Ayra WSM Black 01 fogger machine that exhaled 0.4 L of air per breath.

### Methods of air sampling

For both parts of the study, an IIR type surgical face mask was used as a sample filter and folded over the hose inlet grip of the vacuum cleaner. Two rubber bands (each ripped around twice) made an airtight seal and prevented the mask from being suctioned into the hose. After the application of the sample filter onto the inlet of the vacuum cleaner, the air inlet circle (of about 2.5 cm in diameter) was marked.

In the HCW part of the study, air sampling was performed at approximately 10 cm distance from the mouth for 2.5 minutes per measurement. During each measurement, HCWs were instructed to inhale and exhale deeply, and cough twice every 30 seconds. After each measurement, the sample filter was removed from the hose inlet and carefully inserted into a plastic sampling bag without touching the sample filter. The hose inlet was cleaned with an alcohol-soaked cloth before and after starting every subsequent measurement.

The investigators used protective clothing, FFP2 masks, and eye protection glasses. Before the start of the study, we performed one pilot experiment with an infected volunteer with a high virus load, in which we used a double face mask on the hose inlet grip of the vacuum cleaner. The outside mask tested positive, whereas the inside mask tested negative. This indicated that the air entering the vacuum cleaner did not contain the virus. As an additional precaution, we used a vacuum cleaner with a HEPA filter on the airflow leaving the vacuum cleaner to prevent the potential spread of the virus into the environment.

### Harvesting of viral RNA from the sample filters and face masks

Both sample filters and face masks (i.e., that were worn by infected HCWs during the household experiments) were analyzed on the presence of SARS-CoV-2. Each infected HCW consecutively wore no mask, a cotton non-medical mask obtained from a large international department store, a surgical mask without medical classification that had poor filtration effectiveness, and a surgical mask type IIR that had an effective particle filter. A mouth-shaped area was marked in front of the mouth on each face mask that participants were wearing during the experiments.

In the medical laboratory the marked circle of the sample filters and the marked mouth shapes of the face maskers that participants wore, were cut out using scissors. Subsequently, these cut-out pieces were inserted into separate tubes with each 3 mL PCR extraction buffer and incubated for 40 minutes at room temperature, while during this period samples were also vortexed four times for one minute. Finally, 500 µL of the extraction was used for RNA extraction using the MagNA Pure Total Nucleic Acid Isolating Large Volume Kit (Roche, Germany).

### RT-qPCR

Original patient samples obtained during routine clinical care were tested on our validated in-house RT-qPCR assay according to the national reference method that was established after international collaboration,^8^ the ELITe InGenius® (Elitech, France) platform,^9^ or the GeneXpert Xpress SARS-CoV-2 PCR assay (Cepheid Inc, Sunnyvale, USA) according to the instructions of the manufacturer. For the study, 100 mL from the sample extraction was analyzed on the presence of SARS-CoV-2 RNA by our in-house RT-qPCR assay.

### Statistical analysis

All data were analyzed using Microsoft Excel and R version 3.3.2 (R Foundation for Statistical Computing). Groups were compared by using parametric or non-parametric tests for continuous variables as appropriate and chi-square test or Fisher’s exact test for categorical variables as appropriate. Values of p that were <0.05 were considered to be statistically significant.

## Results

### Vacuum cleaner characteristics

The volumetric airflow into the vacuum cleaner was 97 m^3^ per hour without sample filter, and 29 m^3^ per hour when the sample filter was applied onto the inlet (i.e., corresponding to 483 L per minute).

The airflow velocity was 0.15 m/s (SD 0.06) at 10 cm distance of the hose inlet, and 0.08 m/s (SD 0.03) at 25 cm distance. During air sampling, suction of the air into the vacuum cleaner did not cause any visible changes in the shape or position of the face masks.

In the visualization experiment, aerosols were visibly suctioned into the vacuum cleaner at 10 cm and 25 cm distance when there was no face mask. In contrast, wearing a face mask resulted in a delayed suction of a part of the aerosols that leaked around the borders of the face mask at 10 cm distance, and only minimally at 25 cm (Figure 1 and video in Appendix).

**Figure 1.**
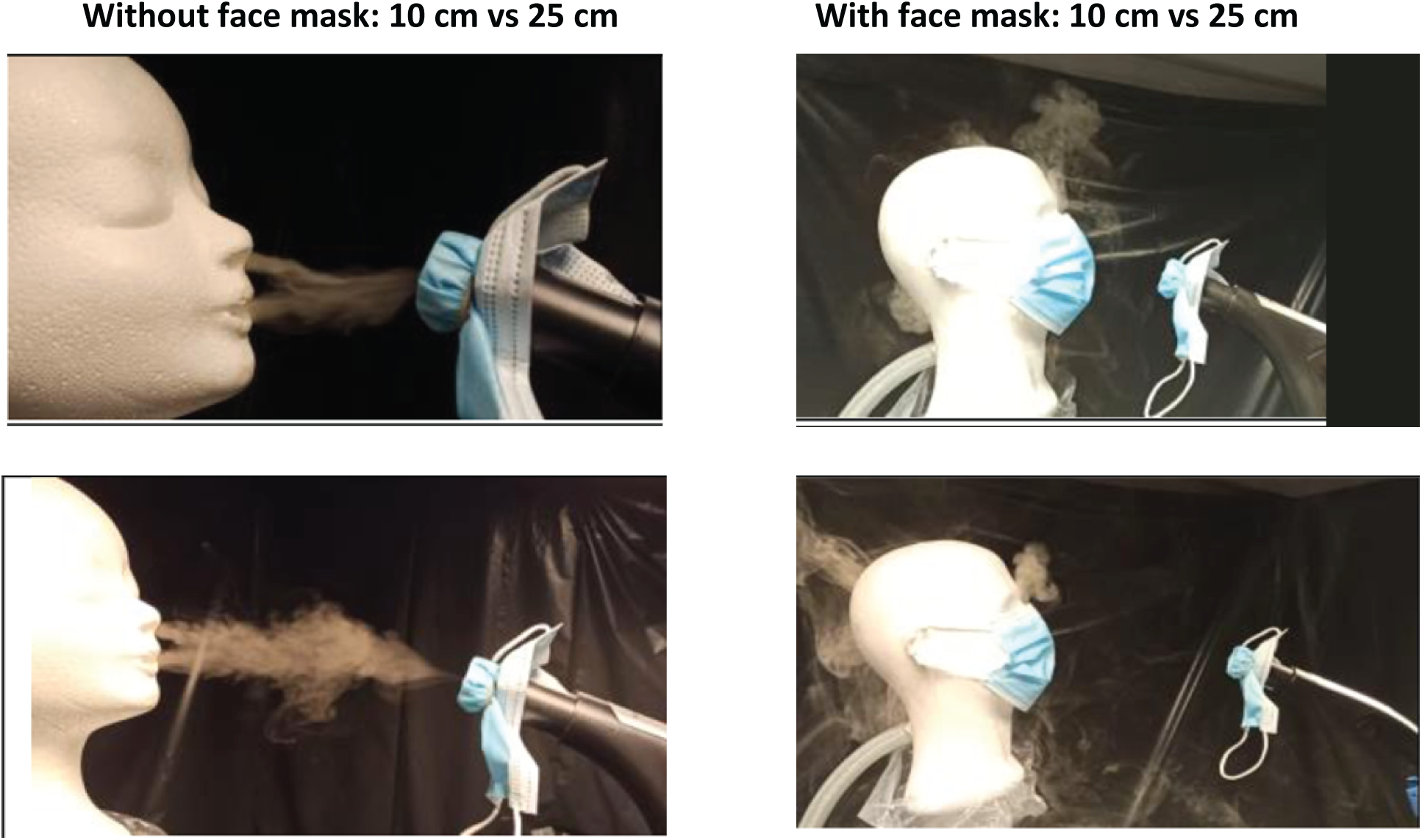
Aerosol visualization experiment. Differences in exhaled fog movements were observed when wearing masks or at different distances between the hose inlet of the vacuum cleaner and the mouth of the infected HCW.

### Observations in HCWs

From 15 HCWs that were screened, 12 agreed to participate and were included within one day after the nasopharyngeal swab sampling date. One HCW was unable because of symptoms related to the infection and two HCWs refused for other reasons. Nasopharyngeal samples of the HCWs had a median Ct value of 17.5 (range 13-19) (Table 1).

**Table 1.** Samples taken in the homes of infected healthcare workers and in hospital rooms of patients.

| Sample origin | Total number tested | Numbers tested positive (%) | Ct value of positive samples – median (range) | Days between nasopharyngeal patient swab and air samples - median (range) |
| --- | --- | --- | --- | --- |
| **12 healthcare workers with recent infection** |  |  |  | 1 (1-2) |
| Nasopharyngeal swab | 12 | 12 (100%) | 17.5 (13-19) |  |
| Air sample in front of mouth: |  |  |  |  |
| uncovered | 12 | 10 (83%) | 35 (32-36) |  |
| covered with cotton mask | 12 | 9 (75%) | 33.5 (32-35) |  |
| covered surgical mask | 12 | 6 (50%) | 35 (32-36) |  |
| covered surgical mask R II | 12 | 10 (83%) | 34 (33-36) |  |
| Free air from room | 5 | 4 (80%) | 34 (32-36) |  |
| Mask sample that patient wore during experiment |  |  |  |  |
| Cotton mask | 12 | 9 (75%) | 33 (29-34) |  |
| Surgical mask <sup>a</sup> | 11 | 6 (54%) | 32 (28-34) |  |
| Surgical mask R II | 12 | 10 (83%) | 33 (30-35) |  |
| **17 intensive care unit patients** |  |  |  |  |
| 6 patients during mechanical ventilation |  |  |  | 4 (0-9) |
| Nasopharyngeal swab <sup>b</sup> | 4 | 4 (100%) | 22.5 (16-26) |  |
| Air sample at 50 cm distance from mouth | 6 | 1 (16%) | 35 (35-35) |  |
| 5 patients during high-flow nasal canulae |  |  |  | 4 (1-5) |
| Nasopharyngeal swab | 5 | 5 (100%) | 28 (17-30) |  |
| Air sample at 50 cm distance from mouth | 5 | 1 (20%) | 33.5 (33-34) |  |
| 6 patients during intubation procedure |  |  |  | 3 (0-6) |
| Nasopharyngeal swab | 6 | 6 (100%) | 28.5 (21-40) |  |
| Air sample at 50 cm distance from mouth | 6 | 2 (33%) | 35 (35-35) |  |

<sup>a</sup> One mask was missing
<sup>b</sup> Two nasopharyngeal swabs were taken in another health care institution

All household visits were performed in the afternoon. HCWs were quarantined in their homes prior to our visit, and some of the subjects were quarantining together with family members who also experienced COVID-19 symptoms. Although the ventilation rate was not measured, windows were closed as the sampling was performed during autumn and winter and mechanical room ventilation was absent.

During the experiments, all HCWs suffered from a dry cough, but no sneezing, no productive cough, or no other “wet” symptoms. Moreover, 10 HCWs had only mild symptoms, with no shortness of breath or dyspnea. Two HCWs experienced shortness of breath following our breathing instructions during the experiment.

Air samples taken in front of the uncovered faces of HCWs were positive in 10 out of 12 (83%) subjects (Table 1). No large droplets were observed on the sample filters after each experiment. The proportion of positive air sample filters was not different between wearing or not wearing face masks: 25 of 36 (69.4%) versus 10 of 12 (83%) (p=0.35), respectively. The face masks that were used by the HCWs were positive in 54% to 83% of cases.

Because of the small differences in viral RNA load retrieved from air samples taken in front of uncovered faces as compared to those in front of the faces that were covered by the face masks during the first experiments, we additionally collected free air samples in the homes (i.e., not near the infected person) at the end of our household visit, and 4 out of 5 samples were positive for SARS-CoV-2 RNA (Table 1).

### Observations in patient rooms of ICU patients

Air samples were collected in 17 ICU patients with COVID-19: six patients during iMV, five during HFNC (60 L/min), and six during an intubation procedure (Table 1). Air samples were positive in 1 of 6 (16%) cases during iMV, 1 of 5 (20%) during HFNC, and 2 of 6 (33%) during the intubation procedure. Of note, intubations were performed using rapid sequency induction including muscle relaxants; all procedures were uncomplicated.

The proportion of positive air samples was lower for ICU patients in comparison to HCWs: 4/17 (%) versus 29/41 (%) (p<0.01); for this comparison we excluded the air samples that were taken in front of the uncovered mouths of the infected HCWs in order to exclude the possible contribution of large droplets.

## Discussion

In this study, we were able to detect SARS-CoV-2 RNA in air samples using a household vacuum cleaner and a routine RT-qPCR test. The presented air sampler method used commonly available material and techniques and was easy to perform. Air samples that were taken in the household environment of recently infected persons carrying high virus loads more frequently contained SARS-CoV-2 RNA in comparison to air samples from patient rooms of infected ICU patients during potential AGMP.

In comparison to other air samplers, our approach has the advantage of including much higher air volumes in order to increase sensitivity, which could explain why many other studies were less successful in detecting SARS-CoV-2 RNA in air samples.^6^ Most currently available air sampling techniques comprise of ‘high-velocity’ impingers which suck airborne virus from the air into a bubbling liquid virus culture medium. These air sampling devices create high shear forces and intense mixing at the air-liquid interface, which may damage viral surface proteins and prevent them from growing in the culture.^3^ This may result in an underestimation of the amount of retrieved viable airborne virus,^10^ and could be one of the reasons that only a few studies have shown the presence of SARS-CoV-2 in air samples.^11,12^

Other possible explanations for the high frequency of positive air samples in the household environment could be related to the specific selection of individuals with high virus loads in a very early phase of the disease and the setting with poor ventilation in which these samples were taken. In poorly ventilated spaces, exhaled aerosols can built-up in the space, creating a higher concentration of possibly infected aerosols.^1,13^ A laboratory study reported that these infectious aerosols can remain viable in the air for up to 3 hours.^14^ The apparently limited effects of different face masks worn by the HCWs could be largely attributed to the study design in which air samples were taken in rooms where the HCWs already spent a number of hours before the test. Assuming that the homes did not have (adequate) ventilation, contaminated aerosols exhaled before the face mask test could have contributed to the high number of positive air samples. This theory was supported by the finding that most free air samples taken not near the patient after the mask experiments were also positive.

Furthermore, SARS-CoV-2 RNA was less frequently detected in air samples obtained in the ICU during AGMP in comparison to the experimental household setting, which is surprising as the risk is deemed high especially during AGMP.^15,16^ These results could be related to the lower virus loads in ICU patients who presented themselves in a later phase of the disease.^17,18^ Moreover, the presence of adequate ventilation in hospital rooms in contrast to poor ventilation in private homes likely contributed to this observation. This is also in accordance with other observations during this pandemic that many infections are more frequently acquired in households in comparison to ICUs or hospitals in general.^19,20^

Although aerosols may leak around the mask borders, excreted large droplets are most likely effectively caught by the masks worn by an infected individual as the majority of masks were positive. Furthermore, we did not observe droplets on the sample filters on the hose inlet of the vacuum cleaner. Also, differences in RNA positivity between settings without wearing a mask and different types of masks were very limited. Together with the aerosol visualization findings in which no exhaled fog appeared to go through the face masks (but only partially around the mask), we postulate that circulating SARS-CoV-2 RNA due to poor ventilation around persons with high virus loads is a plausible explanation for our findings. We consider large droplets to be an unlikely explanation for the positive free air samples or positive air samples taken in front of the mask covered faces. Therefore, it is likely that both the air passing around the mask and aerosols still floating around from the period prior to the actual mask experiment contributed to many positive air samples.

Several studies have shown that different face masks can allow for different levels of leakage, including a previous study that tested fourteen different masks, including surgical masks, KN95, cotton masks, and homemade masks, among others.^21^ The study showed that a tight fit is important to avoid outward leakage through the perimeter, as well as size in general. Similarly, further studies have shown that medical masks stop the forward motion of jets, whether of coughs or breathing, by reducing the speed and redirecting backward, while well-fitted homemade masks with several layers can also reduce the leakage.^22,23^

There are several study limitations to consider. First, the primary aim was to measure the protective effects of different types of face masks worn by infected persons to prevent further spread. In retrospect, our approach failed to address this research question due to our sampling setting with poor ventilation resulting in many positive air samples due to circulating virus RNA. Therefore, our findings should not be interpreted as a failure in the protective effects of face masks. Importantly, SARS-CoV-2 was also detected on the masks worn by the infected persons and thus these masks limited the exposition of the virus to the environment. Second, all our observations were carried out prior to the emergence of variants of concern, such as the United Kingdom, South African, and Brazilian P1 SARS-CoV-2 variants in the Netherlands. We cannot exclude that different results would have been obtained due to more transmittable variants. Third, our experimental findings in adequately ventilated ICU rooms of COVID-19 patients with low proportions of positive air samples during AGMP require confirmation in a larger cohort that should also include different oxygen therapy.

In conclusion, the presented vacuum cleaner based air sampling followed by RT-qPCR is simple to perform, does not require expensive materials, and is an effective method to detect SARS-CoV-2 in the air. Although airborne transmission of SARS-CoV-2 has not yet been widely accepted, it should be considered as an important transmission route, mainly in settings with poor ventilation such as private homes and during an early phase of infection.

## Data Availability

Data are available upon request.

## Acknowledgments

We would like to thank our laboratory technicians, in particular Han Veltman, Gerda Doejaaren, and Dick Wille, and team managers for their assistance in performing the experiments. We would also like to thank Dr. Diana Verboom for her assistance during the conceptual phase of the study.

## Appendix legends

Video material of exhaled fog movements during experiments with and without wearing masks and at different distances between the hose inlet of the vacuum cleaner and the mouth of the mannequin exhaling the fog.

